# Large Language Models Struggle to Encode Medical Concepts — A Multilingual Benchmarking and Comparative Analysis

**DOI:** 10.1101/2025.01.15.25320579

**Authors:** Hossein Rouhizadeh, Anthony Yazdani, Boya Zhang, David Vicente Alvarez, Matthias Hüser, Alexandre Vanobberghen, Rui Yang, Irene Li, Andreas Walter, Douglas Teodoro

## Abstract

Interoperability in health information systems is crucial for accurate data exchange across environments such as electronic health records, clinical notes, and medical research. The main challenge arises from the wide variation in biomedical concepts, their representation across different systems and languages, and the limited context, complicating data integration and standardization. Inspired by recent advances in large language models (LLMs), this study explores their potential role as biomedical knowledge engineers to (semi-)automate multilingual biomedical concept normalization, a key task for semantic interoperability of medical concepts. We developed a novel multilingual dataset comprising 59’104 unique terms mapped to 27’280 distinct biomedical concepts, designed to assess language model performance across this task within five European languages: English, French, German, Spanish, and Turkish. We then proposed a multi-stage pipeline based on a retrieve-then-rerank approach using sparse and dense retrievers, rerankers, and fusion approaches, leveraging discriminative and generative LLMs, with a predefined primary knowledge organization system. Our experiments show that the best discriminative model, e5, achieves an accuracy of 71%, surpassing the best generative model, Mistral, by 2% (p-value < 0.001). For semi-automated workflows, e5 maintained superior performance with 82% recall@10 versus Mistral’s 78%. Our findings demonstrate a pathway to how LLM-based approaches can advance the normalization of multilingual biomedical terms as well as the limitations of LLMs in encoding biomedical concepts.

## 1. Introduction

Semantic interoperability has been sought for many years as an enabler of digital medicine [1, 2]. It ensures that medical terms stored in electronic health records (EHRs) encode a common meaning and can be shared across different health information systems [3, 4]. Despite the importance of semantic interoperability and manual efforts to encode terms using biomedical knowledge organization systems, such as terminologies and ontologies [5, 6], health information systems remain largely un-interoperable [2]. A key challenge with semantic interoperability is the vast number of biomedical entities, making manual mapping or encoding medical terms a highly demanding task. For example, the Systematized Nomenclature in Medicine - Medical Terms (SNOMED-CT) contains more than 350’000 concepts and 1.4 million relationships [7, 8], similar to or exceeding the number of words in many natural languages. Moreover, it is often the case when biomedical terms should retain consistent meanings across different languages. This is particularly important in applications such as personal electronic health records [9, 10], infectious disease surveillance (e.g., COVID-19, antimicrobial resistance) [11, 12], and multi-centric clinical research [13], where health data needs to be accessed or shared across different health settings spanning multiple languages. In these situations, it not only requires domain experts with knowledge engineering skills but also that they are available in multiple languages.

To support the encoding of medical terms and foster semantic interoperability, natural language processing (NLP) methods have been proposed to map free text into concepts defined by biomedical terminologies and ontologies [8]. NLP methods, especially through *information extraction* techniques, can encode medical terms predominantly available in unstructured, free-text format in health information systems into a structured form [14, 15] with a shared meaning. In particular, *biomedical concept normalization* (BCN) or *entity disambiguation* is an NLP task focusing on associating terms within an implicit or explicit context with their most relevant concepts in a knowledge organization system [16]. Early BCN approaches relied on string-matching and dictionary look-up techniques, with tools like MetaMap [17] and cTAKES [18] being widely used. These dictionary-based methods provided interpretability. However, they are heavily lexicon-dependent, hindering their application to multiple languages. Indeed, most of the rules-based BCN tools are only available in English [8].

BCN systems have evolved from rule-based methodologies to advanced machine learning models [19] with improved performance [20]. The application of machine learning to BCN began with models such as Dnorm [21] and TaggerOne [22]. In the last decade, the rise of deep neural networks, including convolutional neural networks [23] and long short-term memory [24] architectures, has facilitated advancements in BCN [25-27]. Recently, transformer-based language models, such as BERT [28] and RoBERTa [29], have demonstrated promising results in BCN, similar to their impact on other NLP tasks across various domains. Language models pre-trained on biomedical and/or clinical corpora, such as BioBERT [30] and Clinical-BERT [31], have been used as the main backbone of several methodologies in this domain [26, 32-36]. Due to the vast size of biomedical terminologies and ontologies, a common machine learning approach for BCN is retrieve-then-rerank, which comprises a two-stage process [37]. Initially, a retrieval component selects the most relevant candidates from an extensive predefined knowledge base of biomedical concepts. Next, a machine learning model reranks the retrieved candidates according to their likelihood of being the correct match for the input term.

While several studies have utilized LLMs in concept normalization pipelines, there has yet to be a study directly comparing discriminative and generative LLMs in the multilingual BCN (MBCN) task. LLMs have demonstrated high capabilities across NLP tasks, generating interest in their potential to automate complex processes that traditionally require domain expertise, such as semantic interoperability. However, their ability to fully meet the demands of complex tasks such as MBCN—especially in multilingual contexts and scenarios with limited term-specific information, such as those in operational EHR databases—remains uncertain. This study addresses this gap by performing a comparative analysis of these two LLM paradigms within the framework of MBCN, focusing on terms with limited context, such as those found in operational EHR databases. While [38] proposed the xMEN system for BCN across multiple languages, their methodology is not comparable to ours as it normalizes the entities by considering their surrounding context. In the proposed MBCN pipeline, we employ several discriminative and generative LLMs to serve as various components, functioning as dense retrievers and rerankers. Utilizing a large-scale dataset comprising 59’104 unique terms across multiple languages and semantic types, we evaluate the performance of various state-of-the-art models within the proposed MBCN pipeline. Our main contributions can be summarized as follows:

1. Inspired by recent advances in information retrieval, we propose a modular MBCN pipeline in which input terms are considered as queries and target concepts as documents. The integrated pipeline combines the power of sparse and dense retrievers, rerankers as well as fusion approaches to normalize multilingual terms across many target ontologies in the biomedical domain.
2. We compare the performance of LLMs, serving as dense retrievers and rerankers, and show how concept normalization effectiveness varies according to LLM type, language, and semantic target. We report results for automated and semi-automated normalization scenarios, demonstrating how combining sparse and dense models and fusing ranks can improve performance.
3. We release a novel biomedical benchmark dataset – MBCN – containing 59’104 unique terms, 27’280 unique concepts, and 113 semantic types across five languages: English, French, German, Spanish, and Turkish.

## 2. Results

### 2.1 MBCN benchmark dataset

To create the MBCN benchmark dataset, we utilized ten manually annotated source biomedical datasets across five languages - English, French, German, Spanish, and Turkish - covering 113 semantic types (Table 1): Medical Concept Normalization (N2C2) [39], BioCreative V Chemical Disease Relation (BC5CDR) [40], QUAERO [41], Unité Commune de Dispensation (Med-UCD), BRONCO-150 [42], DisTEMIST [43], PharmaCoNER [44], Turkish LOINC Linguistic Variant (TLLV)^1^, XL-BEL [45] and Mantra GSC [46]. To simulate a real-world BCN scenario, where terms stored in EHR without an explicit context need to be mapped to a target concept in a knowledge organization system, we extracted the annotations in the source datasets from their context, and the only (implicit) context associated with a term is the target ontology. Thus, each annotation could be considered a biomedical entity to be normalized, such as an item in an EHR database, and the target ontology, e.g., ICD or ATC, is the context, such as the column in which the item is stored. Then, to ensure consistency across different datasets, we transformed all annotation codes into their corresponding Concept Unique Identifier (CUI) using the mapping information within the Unified Medical Language System (UMLS). Thus, an instance in the BCN consists of a triplet *biomedical term*, *target ontology, semantic group, semantic type,* and *CUI,* in addition to the *source language*. In our experiments, we only leveraged the target ontology as the context of input terms. The entire procedure is illustrated in Figure 1, where we pre-process a source dataset to create the MBCN set. We employed the official partitions of the source datasets whenever possible to generate the training, development, and testing partitions of MBCN. We applied a random split for datasets without pre-defined partitions, allocating 60% for training, 20% for development, and 20% for testing.

**Figure 1:**
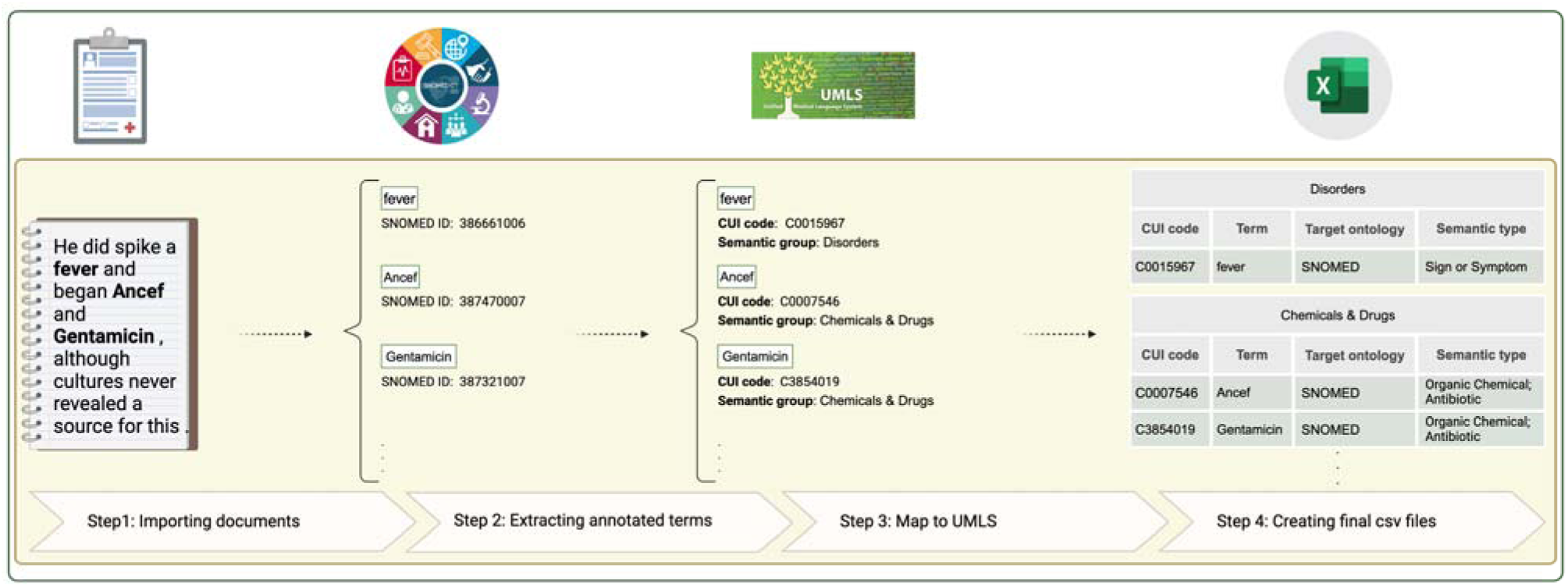
Overview of the MBCN dataset creation process, using an example in which terms are first mapped to SNOMED. UMLS mapping information is then utilized to obtain the corresponding CUI codes and semantic types.

**Table 1:**
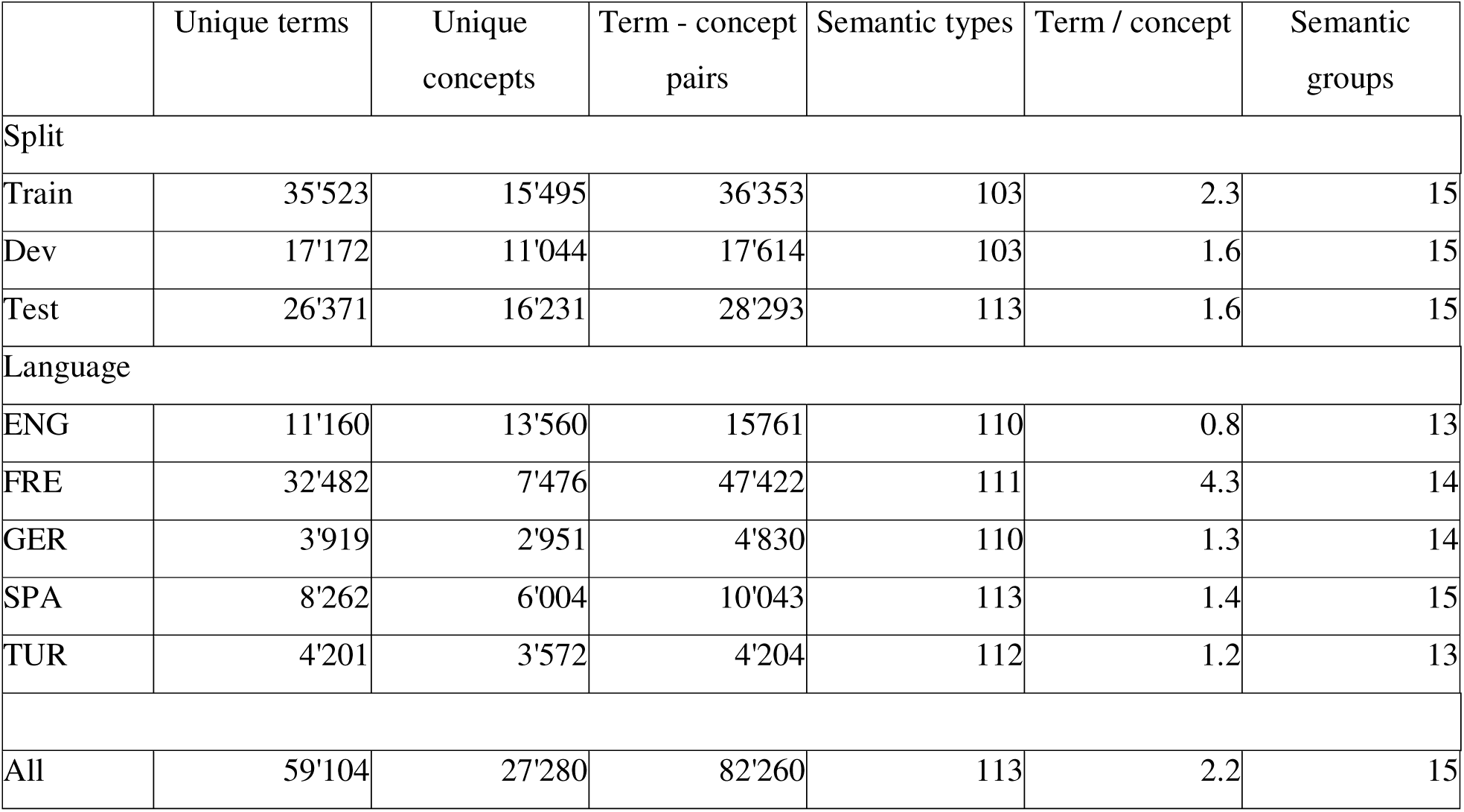
Statistics of the MBCN dataset, stratified by languages and train, dev, and test splits.

|  | Unique terms | Unique concepts | Term - concept pairs | Semantic types | Term / concept | Semantic groups |
| --- | --- | --- | --- | --- | --- | --- |
| Split |  |  |  |  |  |  |
| Train | 35'523 | 15'495 | 36'353 | 103 | 2.3 | 15 |
| Dev | 17'172 | 11'044 | 17'614 | 103 | 1.6 | 15 |
| Test | 26'371 | 16'231 | 28'293 | 113 | 1.6 | 15 |
| Language |  |  |  |  |  |  |
| ENG | 11'160 | 13'560 | 15'761 | 110 | 0.8 | 13 |
| FRE | 32'482 | 7'476 | 47'422 | 111 | 4.3 | 14 |
| GER | 3'919 | 2'951 | 4'830 | 110 | 1.3 | 14 |
| SPA | 8'262 | 6'004 | 10'043 | 113 | 1.4 | 15 |
| TUR | 4'201 | 3'572 | 4'204 | 112 | 1.2 | 13 |
| All | 59'104 | 27'280 | 82'260 | 113 | 2.2 | 15 |

Table 1 presents an overview of the resulting MBCN dataset for the different languages and splits. The dataset contains 59’104 unique terms mapped to 27’280 unique CUIs across 113 semantic types in 5 languages - English, French, German, Spanish, and Turkish - resulting in 82’260 unique term-concept pairs. Among the languages, MCBN contains the highest number of instances in French (n= 47’422). In contrast, it includes an order of magnitude fewer instances in Turkish (n=4’204), making Turkish the language with the fewest terms in the dataset. Note that in English, the number of concepts exceeds the number of terms because a single term can be mapped to multiple CUIs. This occurs because one code from the source ontology, such as MeSH, can correspond to various CUIs. For example, the term ‘famotidine,’ which was originally mapped to the MeSH code ‘D015738’ in the BC5CDR dataset, is associated with five different CUIs: ‘C0015620’, ‘C0678119’, ‘C0733580’, ‘C0733581’, and ‘C0949471’. Following the original test splits of the datasets, 30% (8’562) of the test terms have exact matches with UMLS and 24% (6’835) with the training data; 46% (12’896) have no exact match. While we report results on the whole test set, for transparency, we also disclosed the results on the exact and non-exact match subsets, the latter being the most interesting for our analysis. The complete statistical details for each dataset are provided in Supplementary Table 1.

Figure 2 illustrates the distribution of knowledge organization systems and semantic groups in the MBCN benchmark dataset. ATC, SNOMED, and UMLS are among the most frequently represented ontologies. Five source knowledge organization systems are semantic group specific - ATC and RxNorm for drugs, MedDRA for adverse drug events, LOINC for laboratory, and ICD-10 for diseases - while three are general thesauri and ontologies - SNOMED-CT, UMLS, and MeSH. Among the semantic groups, the categories *Chemicals & Drugs* (71’242; 61%), *Disorders* (29’193; 25%), and *Living Beings* (3’537; 4%) are the three most represented within the MBCN dataset.

**Figure 2:**
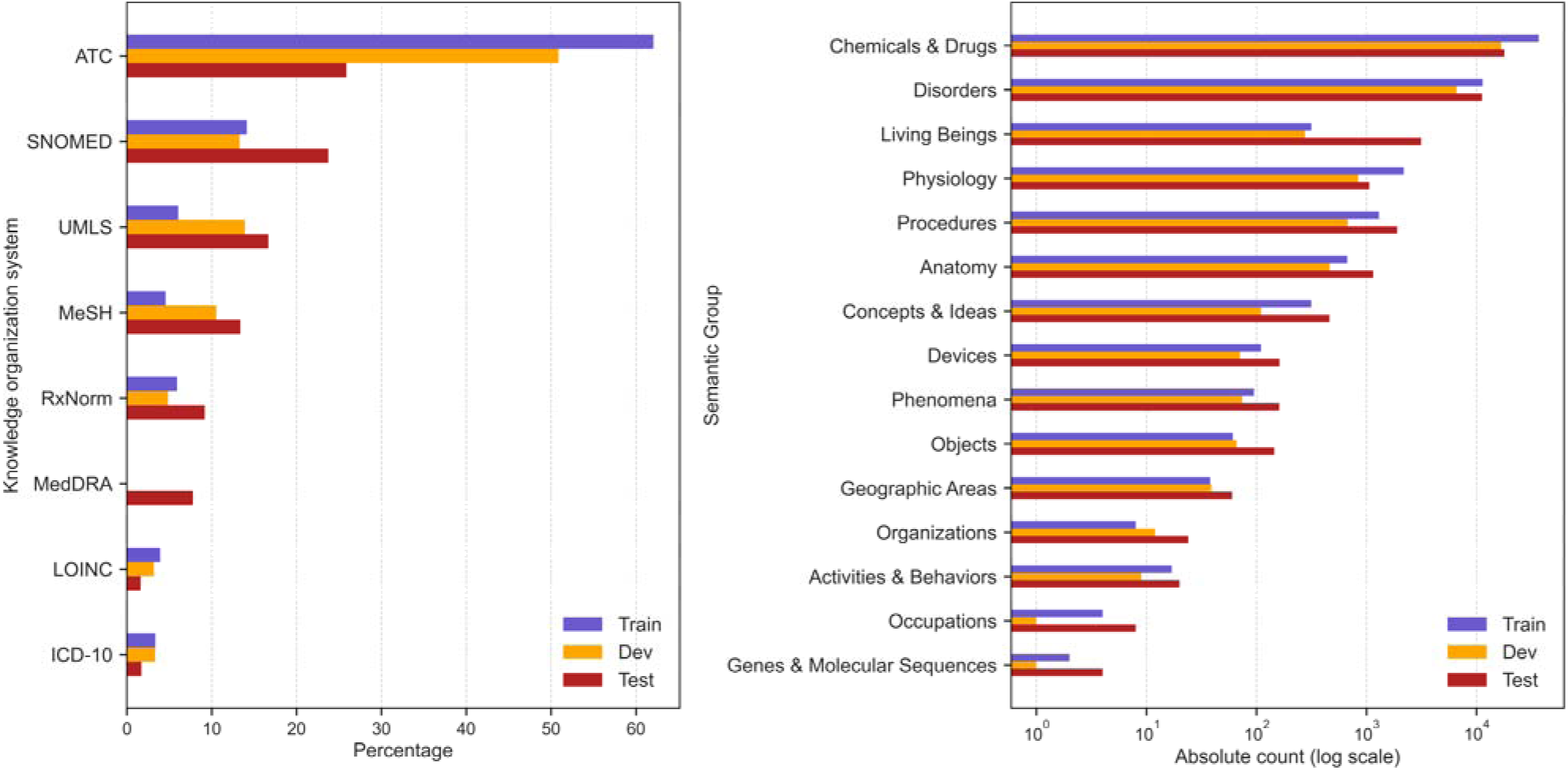
Distribution of concepts within the MBCN benchmark. Left: Knowledge organization systems; Right: Semantic groups

### 2.2 BCN pipeline

The concept normalization task can be formalized as follows. Given a set of terms *P = {p_1_, p_2_, …, p_d_}* to be normalized, e.g., cells from EHR database, and a knowledge organization system KG comprising biomedical concepts *C = {c_1_, c_2_, …, c_n_}*, i.e., the target ontology, the goal is to map each term *p∈P* to its encoding concept *c ∈ C*. To tackle this challenge, we propose a retrieve-then-rerank MBCN methodology, consisting of candidate generation and candidate reranking stages, in which we treat the input term *p* as the query and the set of concepts *C* as the documents. As illustrated in Figure 3, in the initial candidate generation phase, we use a combination of sparse (or lexical) retrieval based on the BM25 [47] framework and dense retrieval techniques based on LLM embeddings to identify the most relevant concepts *c_i_ ∈ C* within the knowledge organization system *KG* (i.e., UMLS) for a given input term *p*. Subsequently, we advance to the candidate reranking stage, leveraging fine-tuned LLMs to evaluate and prioritize the candidates *c*_i_ identified in the previous step. This prioritization is based on the cosine similarity between the vector representations of each candidate and the input term. To leverage the strengths of different models, we use the reciprocal ranking fusion (RRF) [48] algorithm as a rank-fusing technique that combines outputs from various models at each stage. In the Methods section, we provide an in-depth examination of each stage within our proposed framework.

**Figure 3:**
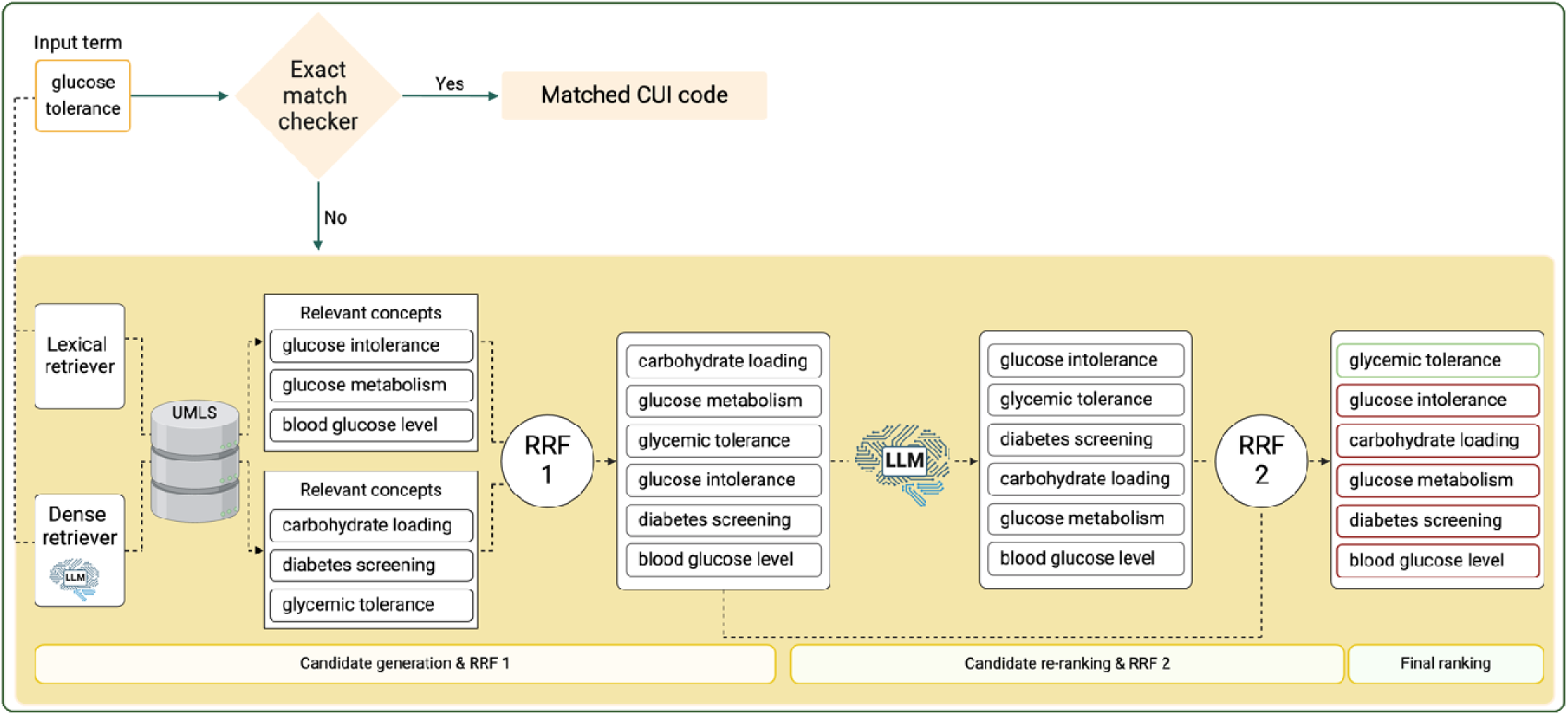
Overview of the MBCN pipeline for biomedical concept normalization. The pipeline receives an input term *glucose tolerance*, which is checked for an exact match within the UMLS knowledge base. As this term doesn’t have an exact match in UMLS, the pipeline advances to the candidate generation stage, where two lexical (BM25-based) and dense (LLM-based) retrieval methods identify relevant concepts from UMLS. An RRF module then merges these two generated ranking lists and produces a new, unified ranking list. In the reranking stage, a fine-tuned LLM computes the cosine similarity between the input term and candidate concepts, followed by a second RRF to determine the final ranking of concepts. As shown, the multistage pipeline successfully brings the correct concept, ‘glycemic tolerance’ (highlighted in the green box), to the top of the ranking list.

Our study examined eight LLMs - four discriminative and four generative - in the dense retrieval and reranking steps of the proposed normalization pipeline. The selection criteria for the LLMs included multilingual training data and open-source availability. The discriminative models were BERT (bert-base-multilingual-cased) [28], DistilBERT (distilbert-base-multilingual-cased) [49], e5 (multilingual-e5-large) [50], and MPNet (paraphrase-multilingual-mpnet-base-v2) [51]. The generative models comprised Aya (aya-23-8B) [52], Mistral (Bio-Mistral-7B) [53], Llama (Meta-Llama-3-8B) [54], and StableLM (stablelm-2-1,6B) [55].

### 2.3 Experiments

#### 2.3.1 Experiment 1

Analyzing the performance of the MBCN pipeline as an automatic concept normalization engine for discriminative and generative LLMs.

This experiment involves stratifying the test instances into two groups: one consisting of terms that precisely match an entry within UMLS and another comprising terms without such direct correspondence. We also report the results that aggregate performance metrics across both instance groups. The performance of different LLMs using the proposed pipeline for automatic MBCN is illustrated in Figure 4. Across the full MBCN dataset, e5 and Mistral models outperform other discriminative and generative LLM alternatives, respectively (p-value < 0.001). The e5 model achieves an overall accuracy of 71%, being 2% above the best generative model (69% accuracy) (p-value < 0.001). Apart from the Llama model, we notice a consistent performance improvement in adding LLMs to the normalization pipeline in comparison with the BM25 baseline (p-value < 0.001). In particular, a performance improvement as high as 6% is seen when adding the e5 model. Interestingly, an important contribution to the model performance is the availability of exact match terms (Figure 4a). We observe a significant performance increase across all language models in all languages, boosting, on average, the best model (e5) performance by 21% (p-value < 0.001). As shown in Supplementary Table 1, 30% of the terms of the MBCN test set appear in UMLS and 24% in their official training split of the source datasets. In terms of language, e5, as the best-performing LLM, achieved its highest effectiveness in French (81%; p-value < 0.001) and English (p-value < 0.001). The model’s performance declined, with accuracy dropping to 59% for Spanish, 50% for German, and as low as 30% for Turkish. These results demonstrate that multilingual normalization remains challenging, even with multilingual LLMs available. The results stratified by source datasets are reported in Supplementary Figure 1.

**Figure 4:**
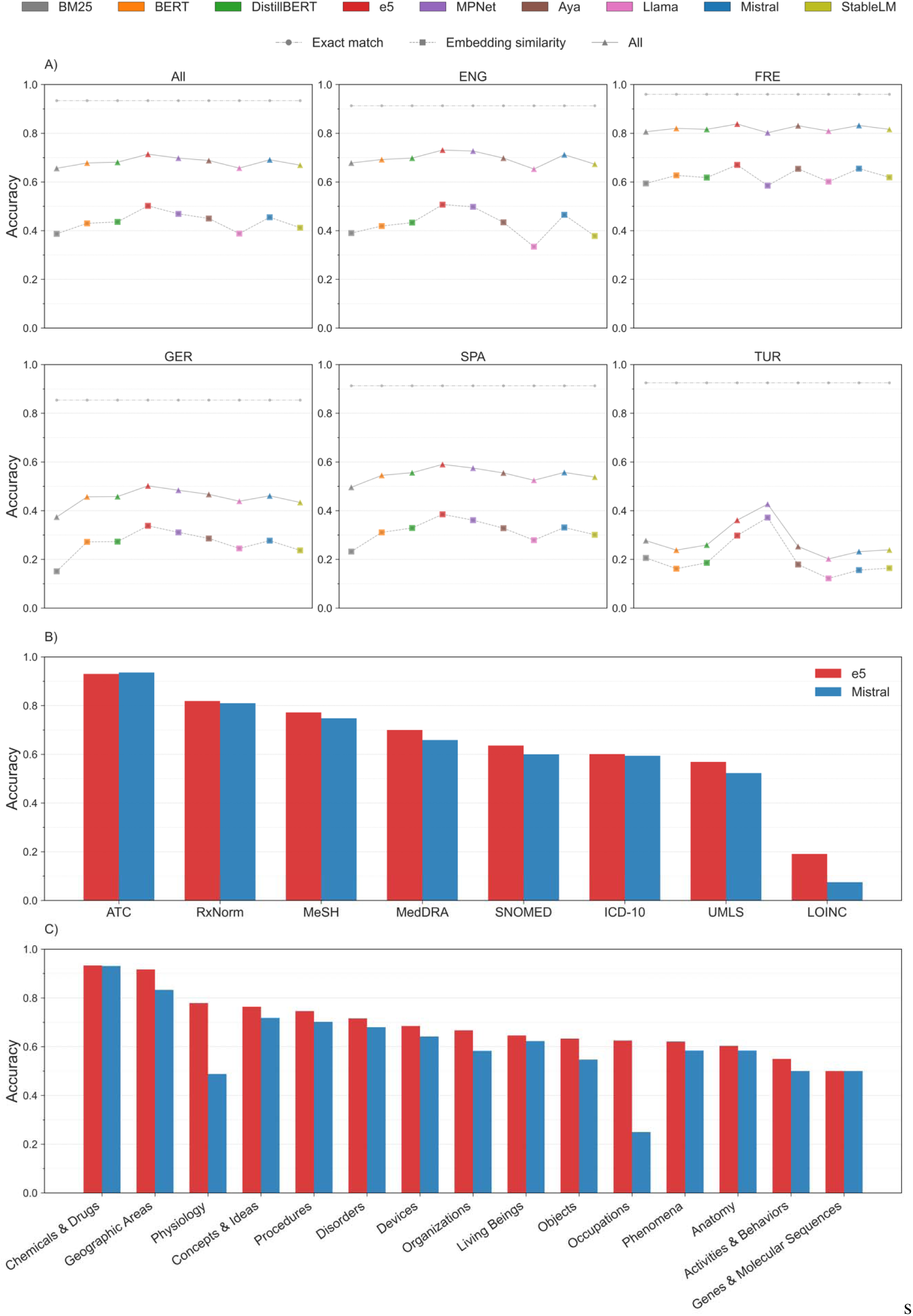
Performance comparison of LLMs on the MBCN dataset stratified by a) language, b) knowledge organization systems, and c) semantic groups.

Regarding target ontology (Figure 4b), the best discriminative and generative models - e5 and Mistral - have mixed results across the knowledge organization systems. Accuracy between 75% and 93% can be seen for ATC, RxNorm, and MeSH, between 57% and 70% for MedDRA, SNOMED-CT, ICD-10, and UMLS, and below 20% for LOINC. ATC was the least challenging knowledge organization system for both models, for which Mistral outperformed e5 (p-value < 0.001), achieving an accuracy of 94% compared to 93% for e5. In contrast, both models performed poorly in linking input terms in Turkish to the LOINC ontology (19% for e5 and 8% for Mistral; p-value < 0.001), indicating that this domain and language present difficulties. Regarding the semantic groups (Figure 4c), e5 demonstrated performance above 80% for *Chemicals & Drugs* and *Geographic Areas*, while performance between 60% and 50% was seen for the *Activities & Behaviors* and *Genes & Molecular Sequences* semantic groups. For the remaining semantic groups, intermediary performance between 58% and 77% was seen.

As illustrated in Supplementary Figure 1, results stratified by the source datasets used to create the MBCN benchmark show variation. Accuracy can go from as low as 23% in the XL-BEL (GER) dataset to as high as 94% in the Med-UCD (FRE) dataset (p-value < 0.001). Aligned with previous results, the best discriminative model, e5, consistently outperformed other models, showing the highest performance in 9 out of 16 source and language datasets (p-value < 0.001). The best generative LLM, Mistral, outperformed the other generative LLMs in 7 out of 16 source and language datasets (p-value < 0.001). However, it did not surpass the performance of the best discriminative model in any of the source or language datasets.

The results of all the language models split by the knowledge organization system are reported in Supplementary Figure 2.

#### 2.3.2 Experiment 2

Analyzing the performance of the MBCN pipeline as a semi-automatic concept normalization engine for discriminative and generative LLMs.

Results in Figure 5a show that the multi-step normalization pipeline - candidate generation (sparse and dense), candidate reranking, and rank fusion - improved the overall performance of the initial sparse model ranking (BM25), with performance generally increasing with each step across all languages. Comparing sparse and dense retrieval methods showed improvements with the dense approaches. For the discriminative LLM, improvements were observed as follows: 10% in recall@1, 16% in recall@3 and recall@5, and 15% in recall@10 (p-value < 0.001). The generative LLM model showed corresponding improvements of 3.1%, 2.4%, 1.1%, and 0.5%, respectively (p-value < 0.001). Additionally, combining sparse and dense retrieval results using reciprocal rank fusion (RRF1) further enhanced performance. For the discriminative model, improvements were observed in recall@10 by 1% in English and 11% in Turkish (p-value < 0.001). The generative model saw improvements across all languages, with a 6% gain in recall@10 (p-value < 0.001). Concept reranking, applied after the first rank fusion step (RRF1) in the normalization pipeline, further enhanced the results. For the discriminative LLM, reranking led to a 2% increase in recall@1 and recall@3 and a 1.5% increase in recall@5. The generative LLM achieved a 1% improvement across recall@1, recall@3, and recall@5. Finally, a secondary rank fusion step (RRF2), which integrated results from RRF1 with those of reranking models, provided additional performance gains. The e5 model achieved average improvements of 0.5%, 1.5%, and 2.1% in recall@1, recall@3, and recall@5, respectively. For the generative LLM, RRF2 improved recall@3 and recall@5 by 1% (p-value < 0.001).

**Figure 5.**
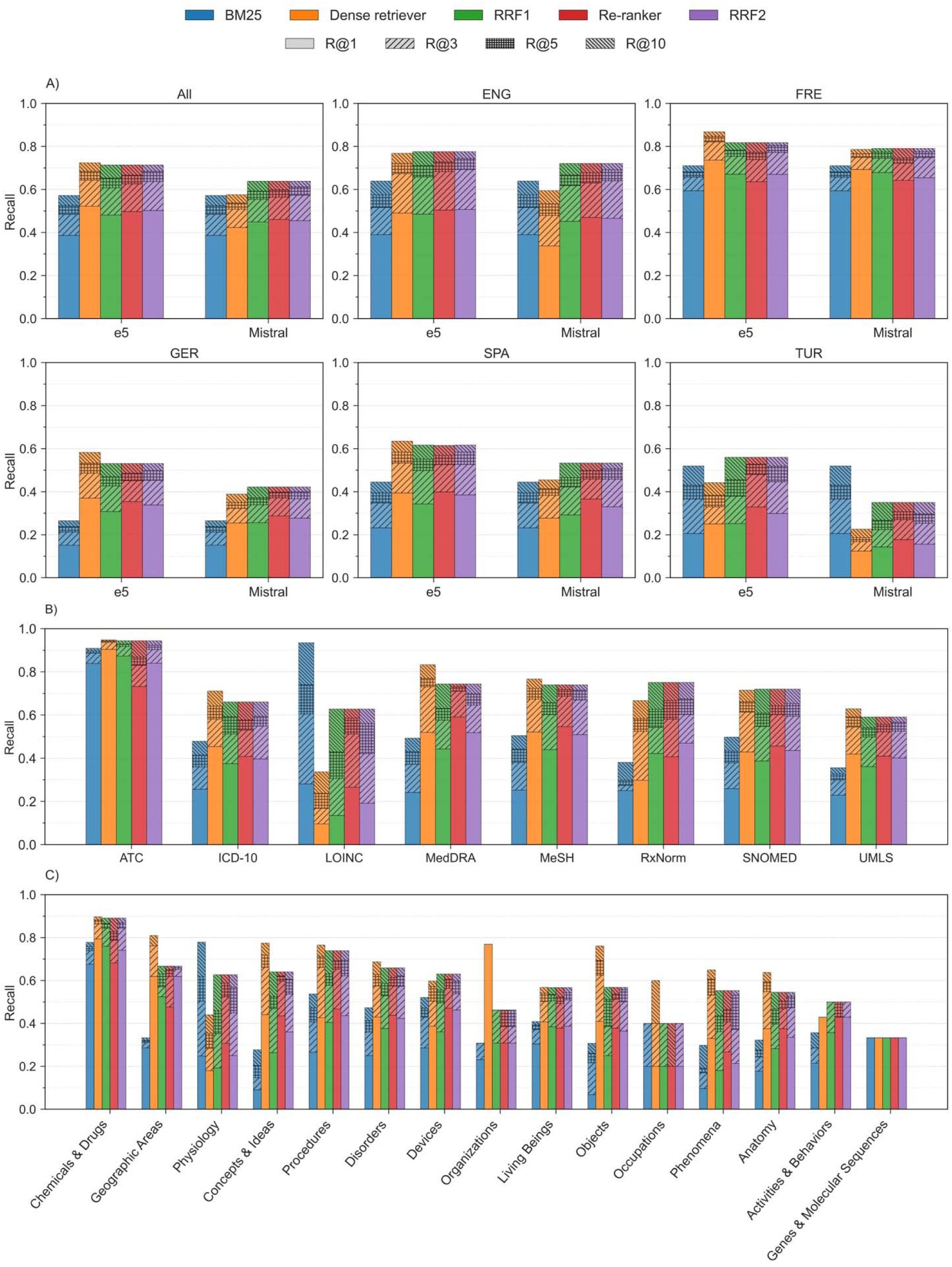
Evaluation of model performance in the concept normalization process across various stages and datasets in a semi-automated scenario. A): Performance of e5 (in left) and Mistral (in right) per language; B): Performance of e5 per knowledge organizations systems. C): Performance of e5 per semantic groups.

Regarding target ontology (Figure 5b), the results reveal that the e5 model demonstrated the most consistent performance when assigning terms to the ATC ontology across all recall metrics (84% recall@1, 90% recall@3, 92% recall@5, and 94% recall@10). Conversely, the model encountered challenges in mapping terms to the UMLS ontology, exhibiting the lowest performance across these recall metrics. For intermediate ontologies, the e5 model’s recall@10 performance ranged between 60% and 75%. A notable finding was the performance of BM25, which uniquely outperformed the e5 model in the LOINC ontology (corresponding to Turkish terms), exceeding it by 60% at recall@10. Examining the semantic groups (Figure 5c), the e5 model showed the performance in the *Chemicals & Drugs* semantic group, achieving a recall@10 of 89%. The *Procedures* semantic group followed with a recall@10 of 74%. In contrast, the *Genes & Molecular Sequences* semantic group proved most challenging for the model, with recall@10 of 33%.

Detailed results, stratified by dataset and language for all tested LLMs, are provided in Supplementary Figure 3.

#### 2.3.3 Experiment 3

Evaluating LLM’s basic understanding of the semantics of multilingual biomedical concepts.

In this experiment, we examine three key aspects of biomedical language understanding by LLMs in a multilingual and restricted context scenario. We hypothesize that LLMs can capture the meaning of biomedical concepts. First, we evaluated overall term-concept semantic similarity using the whole MBCN benchmark. To do so, we considered only the effectiveness of discriminative and generative language models as dense retrievers in the MBCN pipeline. Specifically, we embedded the test instances of the MBCN dataset using LLMs and evaluated the model’s performance in retrieving the relevant concepts corresponding to each input term from the target ontologies using cosine similarity. Second, we evaluated the LLM’s capacity to generate encoding terms in the simplest scenario, i.e., in which terms and concept labels are identical. To do so, we enabled and disabled the exact match checker in the MBCN pipeline and considered only input terms in the test set that had a matching un-capitalized concept label in the knowledge organization system (n=328). Third, we analyzed the impact of case sensitivity on the models’ ability to normalize terms, e.g., input term ‘cancer’ vs. and target concept ‘Cancer’. We identified 190 matching capitalization terms and 138 differing capitalization terms in the previous set and ran the MBCN pipeline with the exact match checker disabled.

##### Term-concept semantic similarity

As shown in Figure 6 (left), the correct concept is ranked in the top position in approximately 90% of the correct concepts found in the top 100 retrieved concepts, followed by an average of 1% distributed across successive intervals of 100 concepts, ranging from 100–200 to 900–1000.

**Figure 6:**
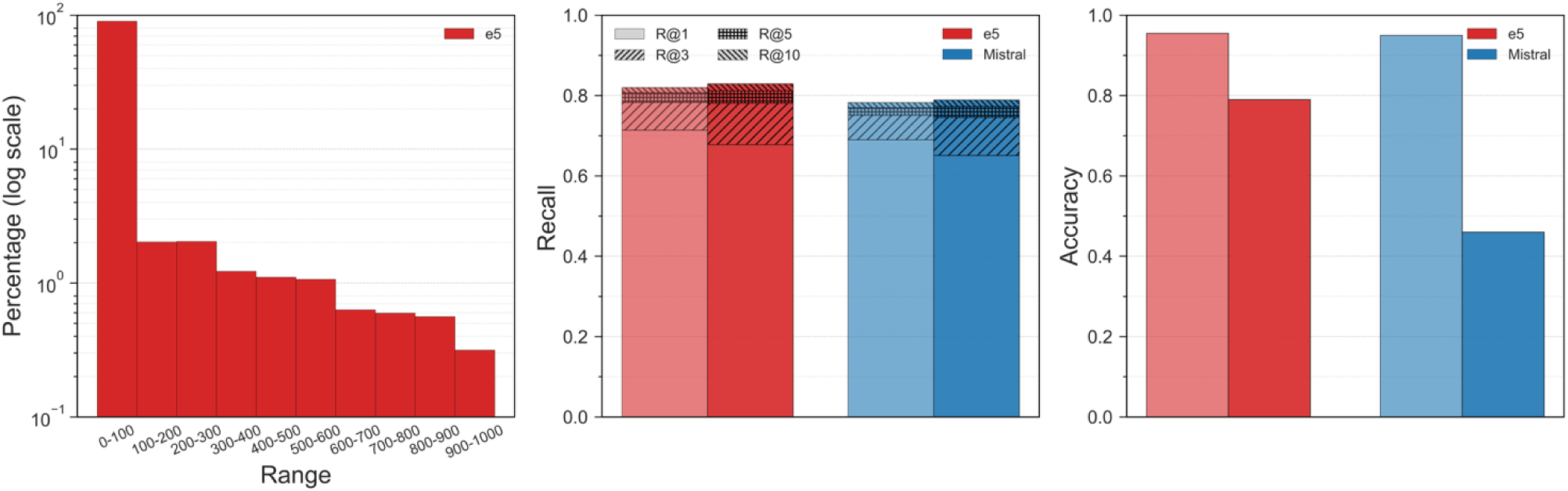
LLMs performance comparison. Left: Impact of exact match checker: light color - exact match disabled, dark color: exact match enabled; Center: Impact of case sensitivity: light color - matching capitalization, dark color: differing capitalization. Right: LLMs as dense retrievers.

##### Same label comparison

As can be seen from Figure 6 (center), enabling the exact match leads to an increase in recall@1 by %5 and 6% for the discriminative and generative LLM, respectively. However, disabling the exact match checker at higher recalls (e.g., recall@5 and recall@10) results in slightly improved performance (approximately 1% for both LLMs).

##### Impact of term’s form on the retrieval accuracy

The results shown in Figure 6 (right) indicate a decrease in accuracy in accuracy for mismatched cases. Specifically, the discriminative LLM experienced a smaller, yet large, performance decline (16%) compared to the generative LLM (50%).

## 3. Discussion

This study evaluated discriminative and generative LLMs for multilingual biomedical concept normalization, focusing on their effectiveness in candidate generation and reranking within a retrieve-then-rerank pipeline. BCN is paramount for semantic interoperability, a long-standing goal in EHR systems, and essential for promoting digital health solutions. However, manual mapping of local EHR terms to knowledge organization systems is costly and requires highly specialized knowledge engineers. This is further challenged in multilingual contexts, for example, in multicentric research and international clinical trials, where normalization of multilingual biomedical concepts is necessary. The development of multilingual LLMs offers opportunities to improve semantic interoperability in healthcare systems.

To assess LLM performance in a limited context scenario, similar to real-world EHR concept normalization, we developed the MBCN benchmark, including 59’104 unique biomedical terms in five languages: English, French, German, Spanish, and Turkish. Moreover, we proposed a methodology inspired by the retrieve-then-rerank information retrieval approach where LLMs are used to rank and rerank matching concepts using semantic similarity in the embedding space and as fine-tuned text classifiers, respectively. The proposed MBCN methodology achieved an accuracy of 71% using a discriminative model (e5), compared to an accuracy of 69% using the best generative model (Mistral). In the semi-automated scenario, focused only on the unseen terms, the best discriminative model achieved a recall@10 of 82%, 4% higher than the best generative model.

Our analysis of LLMs’ semantic understanding of biomedical concepts revealed both strengths and limitations in multilingual contexts. The high concentration (over 90%) of correct concepts ranked in the top 100 retrieved results, (among approximately 4.5 million concepts) demonstrates that LLMs can effectively capture fundamental semantic relationships in biomedical terminology. While this indicates strong initial retrieval capabilities, it also emphasizes the need for developing more precise reranking mechanisms to identify the correct concept within these candidate lists. Furthermore, the impact of surface-level variations, particularly case sensitivity, exposed important vulnerabilities in LLMs. When evaluating case-mismatched terms, we observed accuracy drops of 16% for e5 and 50% for Mistral, indicating that these models are highly sensitive to surface-level text variations for generating embeddings. This finding is particularly relevant for clinical implementations, where variations in text formatting and capitalization are common in EHR systems.

Performance variations between discriminative and generative models could be related to several factors, including training tasks, training data, and parameter size. For example, the better performance of the best discriminative model (e5) may be justified by the pre-training strategy, which uses weak supervision signals from a large, curated text-pair dataset called CCPairs [50]. This contrastive pre-training allows the model to distinguish between synonymous and non-synonymous terms by positioning semantically similar terms closer together in the embedding space. This capability aligns with the primary objective of the concept normalization task: to assign the highest rank to the concept that accurately reflects the meaning of the given input term (i.e., synonym) while ranking non-synonymous concepts lower in the output list.

Regarding generative models, the superior performance of Mistral may be attributed to its pre-training on biomedical corpora, which provides the model with more comprehensive domain-specific knowledge. In contrast, the StableLM model consistently achieves the lowest performance across all evaluated datasets. This underperformance might be explained by StableLM’s relatively smaller number of parameters (1.6 billion) compared to other generative LLMs evaluated in the study (between 7 and 8 billion). However, it is important to note that despite its lower performance, StableLM still has one order of magnitude larger parameter count than the best-performing discriminative model, i.e., e5, with 354 million parameters. This observation suggests that while parameter count can influence model performance, it is not a solid determining factor, especially when comparing different model architectures and training approaches.

The difference in language coverage may contribute to the performance differences between discriminative and generative LLMs. While discriminative models have been trained in over 100 languages, generative models lack such breadth. However, this alone does not explain their underperformance, as even in widely represented languages such as English, generative models still can not beat their discriminative counterparts. This suggests that additional factors, such as training strategies and model architecture, may impact the observed performance differences.

The analysis of the source datasets used to build the MBCN benchmark shows that 30% of the test terms are found in metaknowledge organization systems, such as UMLS. Performance analysis of the exact match checker identifies difficulties in managing abbreviations and terms with limited context. For instance, in terms sourced from the N2C2 dataset, the term ‘PVC’ is misassigned to ‘Polyvinyl Chloride’ instead of ‘Premature Ventricular Contractions’ when using exact matching. Similarly, the abbreviation ‘MI’ is erroneously linked to ‘Mental Institution’ rather than ‘Myocardial Infarction’. These types of errors could have significant clinical implications, potentially affecting patient care decisions if not properly addressed in the implementation phase.

The classic information retrieval model, based on the sparse BM25 framework, demonstrated competitive performance against several cutting-edge LLMs in some scenarios despite its reduced computational complexity. However, a closer examination of BM25’s performance on cases without exact matches in their corresponding ontologies or training sets reveals its limitations. In this scenario, BM25’s performance consistently falls behind the full MBCN pipeline, with an accuracy drop of 12%. Unlike LLMs, which leverage deep contextual embeddings to match concepts without explicit term overlap, BM25 is constrained to surface-level matching based on term frequency and document relevance scoring.

Despite several experiments conducted on the large-scale MBCN benchmark, this study has important limitations that need to be considered for clinical implementation. First, while our study included five languages - representing the largest evaluation of MBCN without context to date - many other languages remain understudied. This could limit the system’s utility in diverse healthcare settings. Second, larger generative LLMs were not fully explored due to computational constraints, though preliminary experiments with Llama-3-70B showed challenges in dense retrieval performance. Finally, our study covered only 27’280 CUIs out of approximately 4.5 million available in the UMLS, which could affect real-world clinical applications.

In conclusion, this study conducts a comparative analysis of several discriminative and generative LLMs, demonstrating their potential for advancing the multilingual biomedical concept normalization. Our findings reveal that a discriminative model, e5, with 345 million parameters, outperforms the generative counterparts containing a magnitude larger parameters (1.6 to 8 billion), indicating the importance of specialized training approaches over model size and scaling. Further analysis shows that LLMs witness a notable drop in performance when dealing with new, unseen terms compared to ones they have previously encountered. In addition, while LLMs demonstrate strong performance in normalizing drug-related terms, they face challenges with other term categories, suggesting opportunities for further enhancement across other areas. A promising direction for future research would be to consider expanding language coverage beyond the current five languages and exploring hybrid architectures that combine the strengths of both discriminative and generative models. As LLM development continues to evolve, we anticipate further improvements in biomedical concept normalization systems. These advances promise to enhance semantic interoperability across healthcare systems while addressing the increasing demand for automated multilingual terminology standardization.

## 4. Materials and methods

### 4.1 Knowledge organization system

We employed UMLS 2022AB as the primary knowledge organization system. It integrates over 4 million biomedical concepts from terminologies such as MeSH, SNOMED CT, and MedDRA. We used UMLS concepts to encode terms in the MBCN benchmark as the knowledge organization system in the MBCN normalization pipeline.

### 4.2 Source datasets used to create the MBCN benchmark

The MBCN benchmark dataset integrates ten publicly available datasets across five languages—English, French, German, Spanish, and Turkish—covering 113 semantic types. For English, we used N2C2 [39] (discharge summaries mapped to SNOMED-CT and RxNorm) and BC5CDR [40] (chemical-disease entities mapped to MeSH). French datasets included QUAERO [41] (biomedical entities from MEDLINE abstracts) and Med-UCD (medication codes linked to ATC). German data are sources from BRONCO-150 [42] (oncology annotations mapped to ATC and ICD-10), while Spanish datasets included DisTEMIST [43] (oncology-related clinical annotations) and PharmaCoNER [44] (chemical and protein entities). For Turkish, we used the TLLV dataset^2^, which maps biomedical Turkish terms to LOINC terminology. We also incorporated two multilingual datasets: XL-BEL [45], spanning English, German, Spanish, and Turkish, and Mantra GSC [46], covering English, French, German, and Spanish, with concepts mapped to SNOMED-CT, MeSH, and MedDRA. Predefined dataset splits were retained where available, and others were divided into training (60%), development (20%), and test (20%) subsets. Supplementary Table 2 summarizes the datasets’ languages, ontologies, entity types, and semantic coverage.

### 4.3 MBCN pipeline

The MBCN pipeline consists of the following steps (depicted in Figure 3):

I. **Input processing:** Receives a biomedical term *p ∈ P* and a knowledge organization system KG containing the target concepts *c ∈ C* as input. UMLS was used as the knowledge organization system KG, and UMLS CUIs defined the set of concepts *C* in our experiments.
II. **Exact match check:** Matches input term *p* to the labels of each concept *c* in the knowledge organization system KG using exact string matching. If no match is found, the process advances to retrieval and reranking steps III to VI.
III. **Candidate generation:** Leverages two retrieval models, sparse and dense, to retrieve potentially relevant concepts *c ∈ C* for the input term *p*.
IV. **Rank fusion (RRF 1):** Combines rankings from sparse and dense retrievers using a ranking fusion algorithm to produce a combined candidate list.
V. **Candidate reranking**: Uses a fine-tuned reranker based on discriminative or generative LLM to rerank the candidates’ list generated in the previous step. The LLM employed in this step is identical to the one utilized for dense retrieval.
VI. **Final rank fusion (RRF 2)**: Combines rankings from steps IV and V to produce the final ranked list of concepts.

During experiments, dataset-specific filtering was applied, such as restricting UMLS concepts to RxNorm and SNOMED terminologies for the N2C2 dataset. Furthermore, we augmented UMLS by integrating training instances of the MCBN dataset, each linked to its corresponding concepts.

#### 4.3.1 Candidate generation

We employed the BM25 as a sparse lexical-based retriever, a probabilistic method that extends the term frequency-inverse document frequency model by accounting for term frequency, document length, and average document length to determine relevance. Additionally, we used eight different multilingual discriminative and generative LLMs as dense retrievers. Our methodology involves first generating embeddings for all labels of the UMLS concepts, using mean pooling representation from the last hidden layer of each LLM. These embeddings are then stored in a vector database to facilitate retrieval processes. For each input term p, we retrieve the top-k most similar set of labels and then associate them to their corresponding concepts *{c_1_, c_2_, c_3_, …, c_k_} ∈ C*. In our experiments, we set k to 1,000 and used Qdrant as the vector database in our implementation

#### 4.3.2 Rank fusion

Reciprocal rank fusion (RRF) [48] is a widely used and effective method for combining ranking results from multiple retrieval systems, merging the strengths of different retrieval systems to improve performance. We used the RRF algorithm to combine ranking sets from different retrieval systems. For each candidate, the RRF score is calculated as:

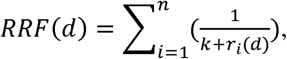

where *RRF(d)* is the final score for document *d*, UMLS concept *c* in our case, *n* is the number of retrieval systems, *r_i_(d)* is the rank of document *d* in the *i_th_* ranking list, and *k* is a constant (set to 60). We apply this algorithm twice within our pipeline: first, to combine the results of BM25 and the dense retriever (RRF1), and second, to merge the outputs of RRF1 with the reranker (RRF2).

#### 4.3.3 Candidate reranking

As the second phase of our retrieve-then-rerank normalization approach, the reranking step employs fine-tuned LLMs, either a discriminative LLM fine-tuned with Sentence-BERT (SBERT) [51] or a generative LLM fine-tuned with LLM2Vec [56] architectures. The details for fine-tuning different types of language models are discussed in Section 4.4. We used the fine-tuned models to rerank the formerly generated ranking list (in step III) based on their semantic similarity to the input term. To do so, we extract the corresponding UMLS labels for each concept *c* in the ranking list and calculate their cosine similarity with the input term. The representative score of *c* is determined by the following formula, i.e., averaging the cosine similarity scores of the top *m* most similar synonyms to the input term:

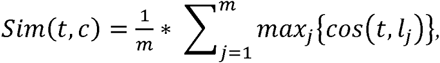

where *sim(t, c)* is the similarity score between term *t* and *c*, and *cos(t, l_j_)* is the cosine similarity between *t* and the *j_th_* label of *c* in UMLS. Finally, concept labels are sorted based on their corresponding scores to create the final ranking list.

### 4.4. Fine-tuning large language models

As the first step of utilizing the language models as a reranker in our normalization pipeline, we created a pair-wise dataset consisting of term-concept pairs via UMLS mapping information. We fine-tuned LLMs using our dataset, employing either SBERT [51] for discriminative fine-tuning or LLM2Vec [56] for generative fine-tuning. The subsequent section explains the fine-tuning process using our constructed dataset and the methodology for leveraging these refined models as rerankers.

#### 4.4.1 Dataset construction

We developed a novel dataset for fine-tuning LLMs, focusing on identifying whether a pair of biomedical passages conveys the same meaning.

The pair generation process involves three key steps (illustrated in Figure 7):

1. **Concept retrieval (Step 1)**: Uses the BM25 retrieval module to extract the top 20 UMLS concepts *{c_1_, c_2_, …,c_20_}* for each source term in the MBCN training set.
2. **Concept ranking (Step 2)**: Re-orders the retrieved concepts, positioning the correct concept at the top (in green), followed by the remaining concepts (in red).
3. **Term-concept pair generation (Steps 3, 4, and 5)**: Generate term-concept pairs using the labels of the retrieved concepts. Using concept *c* in the positive group (in green), we extract all its corresponding UMLS labels and create positive instances either by pairing them with the input term *t* or one another. In the negative group (in red), we generate negative pairs by combining labels from different concepts in the list. For instance, a negative pair such as (*l_2,1_, l_5,3_*) is formed by taking the first label from the second concept and the third label from the fifth concept.

**Figure 7:**
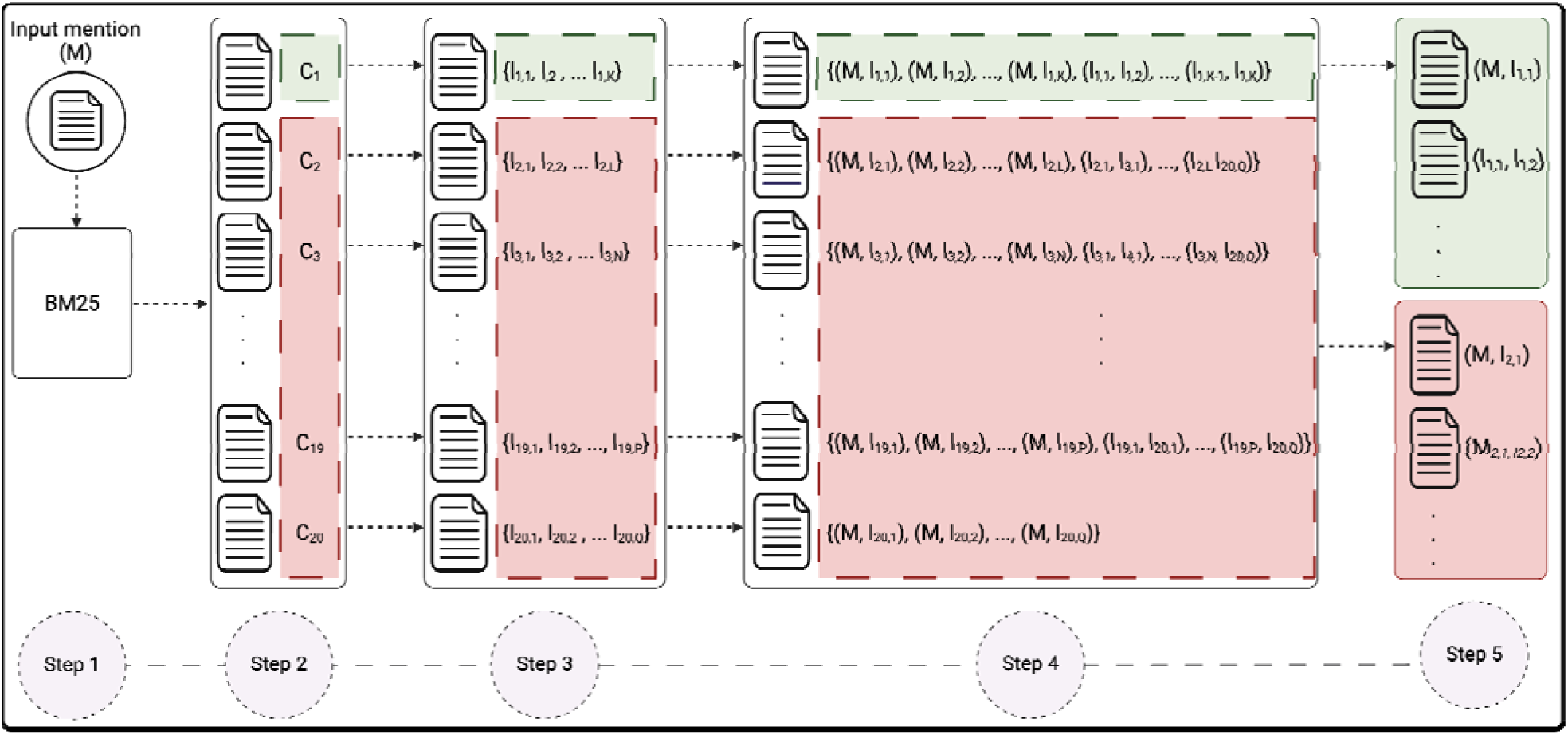
Generating contrastive positive and negative pairs using BM25 search results to create training datasets for language model fine-tuning. Green and red boxes represent the instances for generating positive and negative pairs, respectively.

Through the data generation process, we extracted 2 million monolingual and multilingual biomedical pairs. The dataset was balanced to ensure equal distribution between positive and negative instances. We randomly partitioned the data into training (60%), development (20%), and test (20%) sets, with an equal distribution of positive and negative pairs in each split. Each positive pair was labeled as 1, and each negative pair as 0. These labeled pairs were then used to fine-tune targeted discriminative and generative models using the SBERT and LLM2Vec architectures, respectively.

#### 4.4.2 Fine-tuning details and experimental setup

The computational infrastructure for model development comprised a cluster of eight NVIDIA RTX 3090 (24 GB) GPUs. We utilized the Sentence-BERT-v3 framework to train the discriminative LLMs. Each LLM was trained on the entire training set of our constructed dataset, 1.2 million data points, using a batch size of 8. The fine-tuning process required approximately three days of computational time. To manage computational resource availability, generative LLM fine-tuning was conducted using a reduced dataset of 300’000 paired samples.

Employing the LLM2Vec framework, the fine-tuning process was executed with a batch size of 4 per GPU, with training durations extending up to seven days, depending on the size and architectural complexity of each LLM. Throughout all experimental procedures, we maintained the default parameter configurations provided by the respective frameworks.

### 4.5 Evaluation metric and hyperparameter tunning

We assessed the performance of models in our MBCN pipeline using the recall at k (R@k) metric as our primary evaluation criterion. This metric quantifies the proportion of documents, UMLS CUIs in our case, that appear within the top *k* predictions for each model. Recall@1 is equivalent to ‘Accuracy,’ representing the proportion of instances where the topranked prediction correctly matches the ground truth label.

Hyperparameter optimization focused on two critical aspects of our approach. First, we fine-tuned the BM25 parameters (*b=0.1, k_1_=1*) through dataset-specific optimization. Our experimental process explored two key hyperparameters: (1) the number of concepts input to the reranker for each input and (2) the number of top similar labels within each concept considered during the ranking process (denoted as m in the formula within Section 4.3.3). Through our evaluations of the development set, we identified configurations that performed best. Specifically, we found that inputting 10 concepts to the reranker for each output produced the most effective results. Moreover, setting *m* to 3—the top 3 similar labels of each concept—proved optimal for scoring and ranking the concepts.

## 5. Code and data availability

This study employs two primary datasets: (1) 10 benchmark datasets spanning five languages and (2) data extracted from the UMLS for language model fine-tuning. These benchmark datasets are publicly accessible. However, the N2C2 and BRONCO datasets require explicit access permission from their respective creators. Upon acceptance of the paper, we will provide documentation detailing the data access procedures and methodological instructions for recreating the constructed datasets.

The entire research process, including dataset development and experimental conduct, was implemented using Python. Upon the paper’s acceptance, we will make the complete source code publicly available.

## Author contributions

DT and HR conceptualized the study. HR and DT drafted the initial manuscript. HR, AY, DV, and BZ performed the experiments. All authors read and approved the final version of the article. DT supervised all steps of the work.

## Data Availability

Data will be available.

## Footnotes

1 https://loinc.org/linguistic-variants/

2 https://loinc.org/linguistic-variants/

## Supplementary Information

**Supplementary Table 1:**
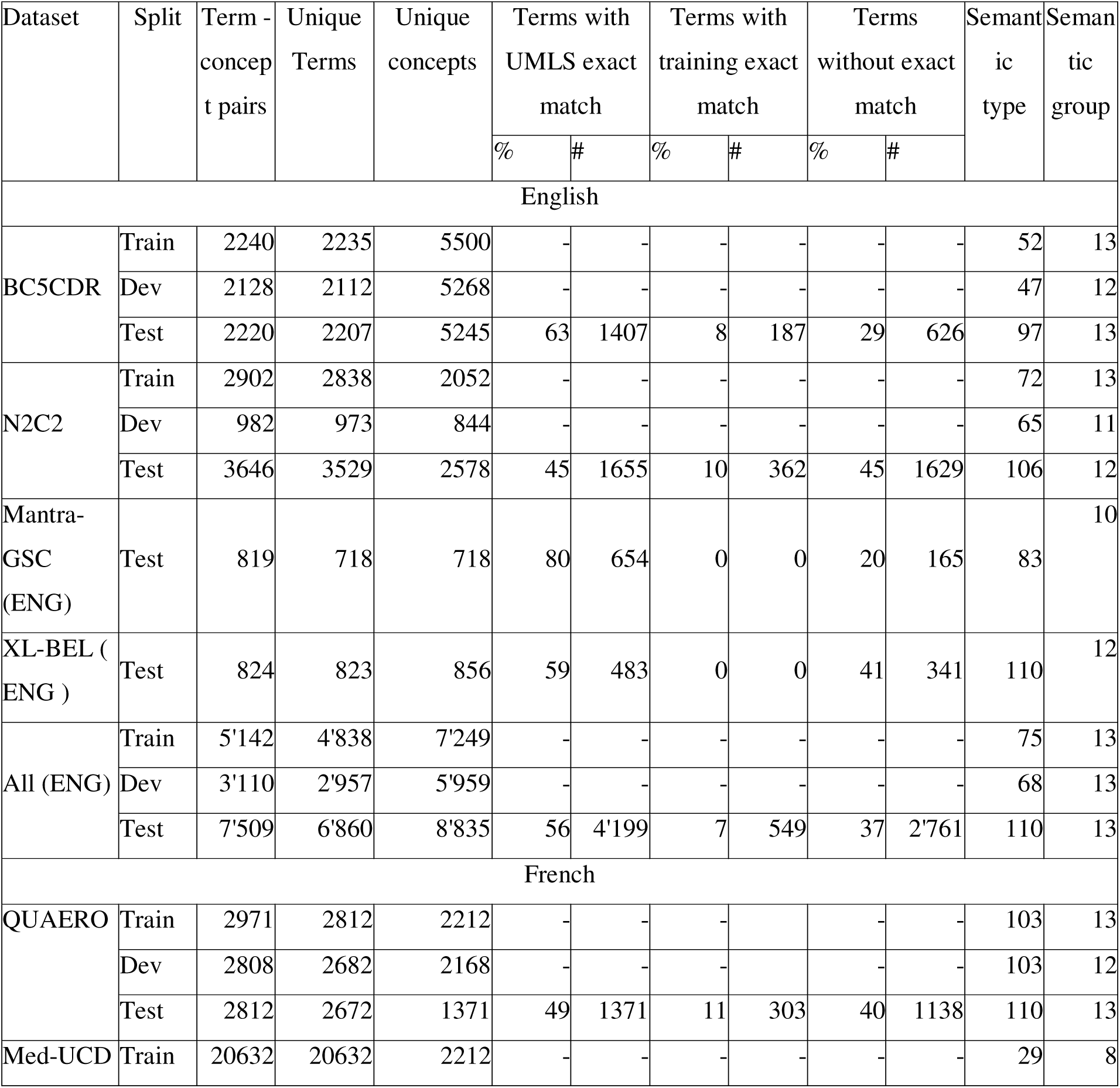
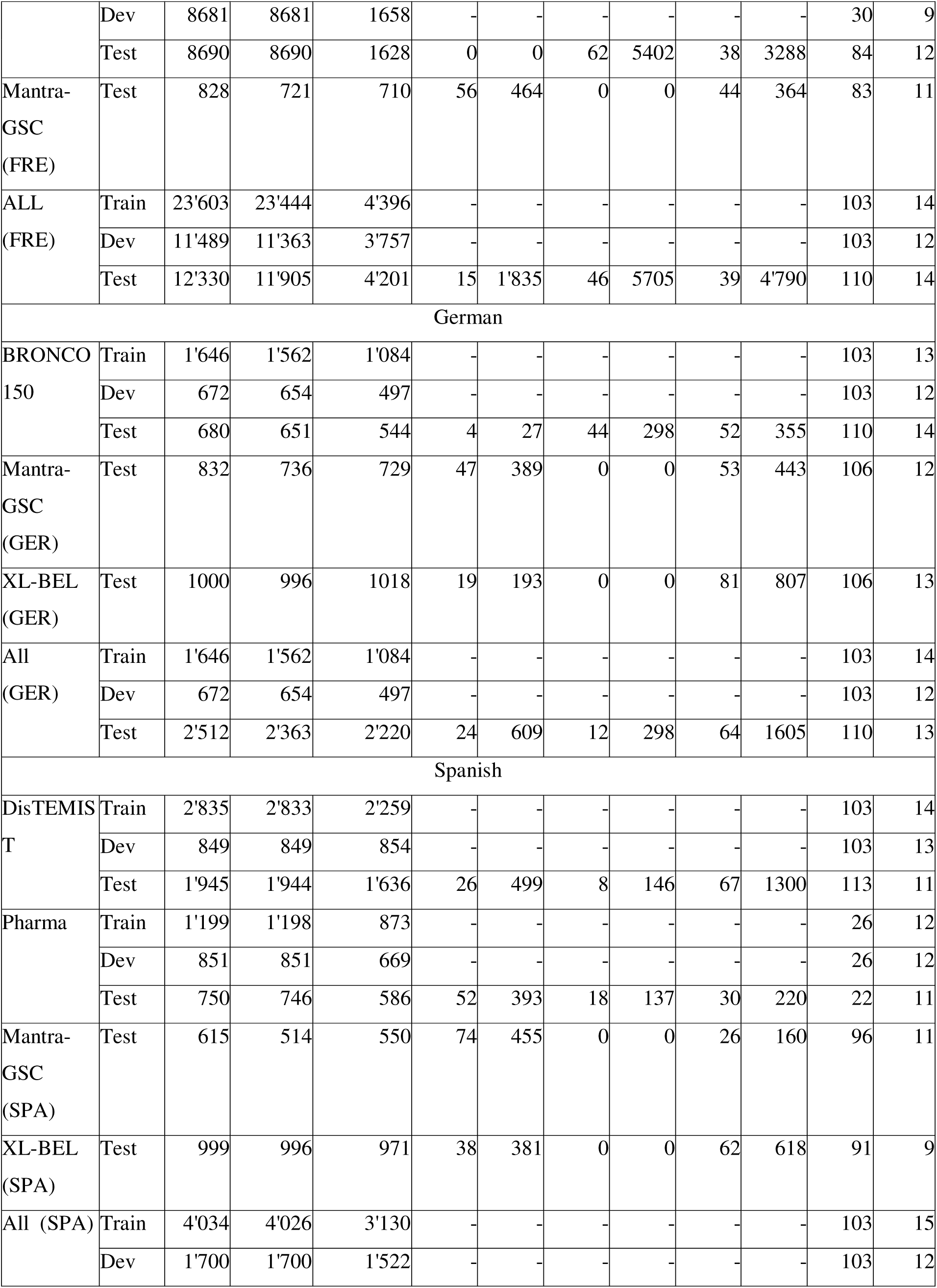
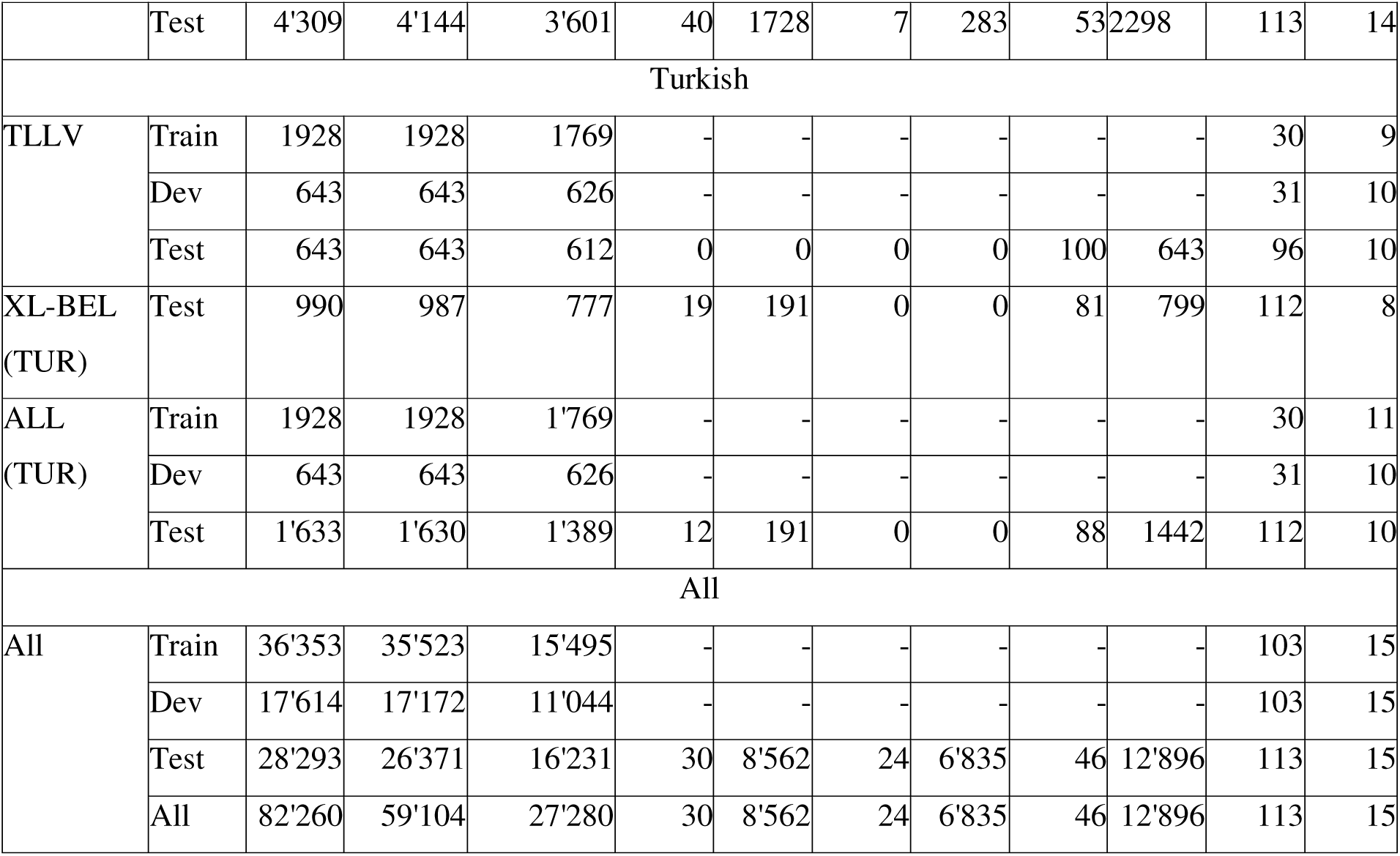
Statistical details of each MBCN dataset, divided by source datasets and languages.

**Supplementary Figure 1:**
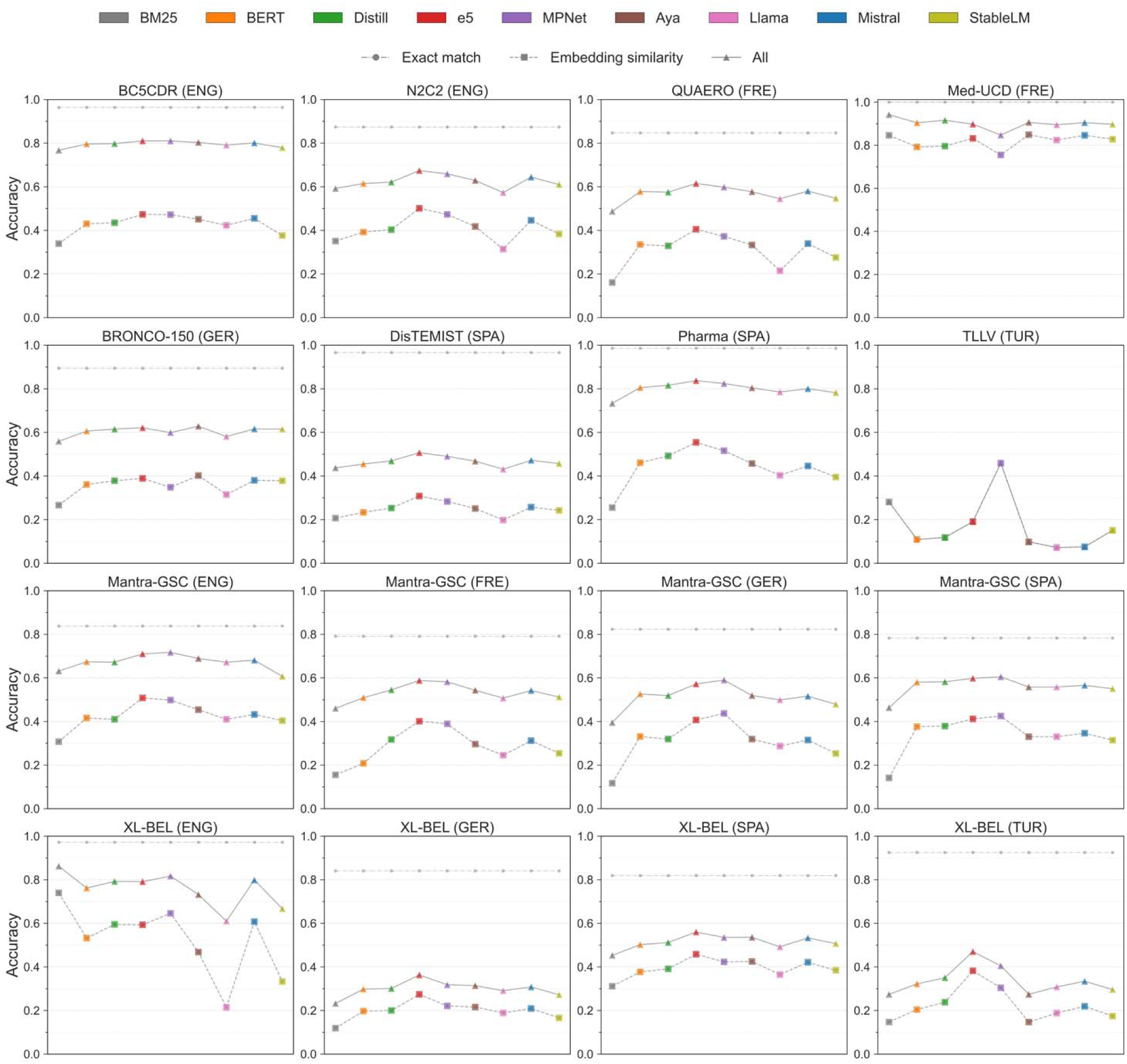
Performance comparison of LLMs on the MBCN dataset, divided by datasets and languages. In each plot, the lines with circles represent the accuracy of the exact match checker on instances of Group A, while the lines with squares and triangles illustrate the performance of LLMs on instances without exact match and their average performance across the entire dataset, respectively.

**Supplementary Figure 2:**
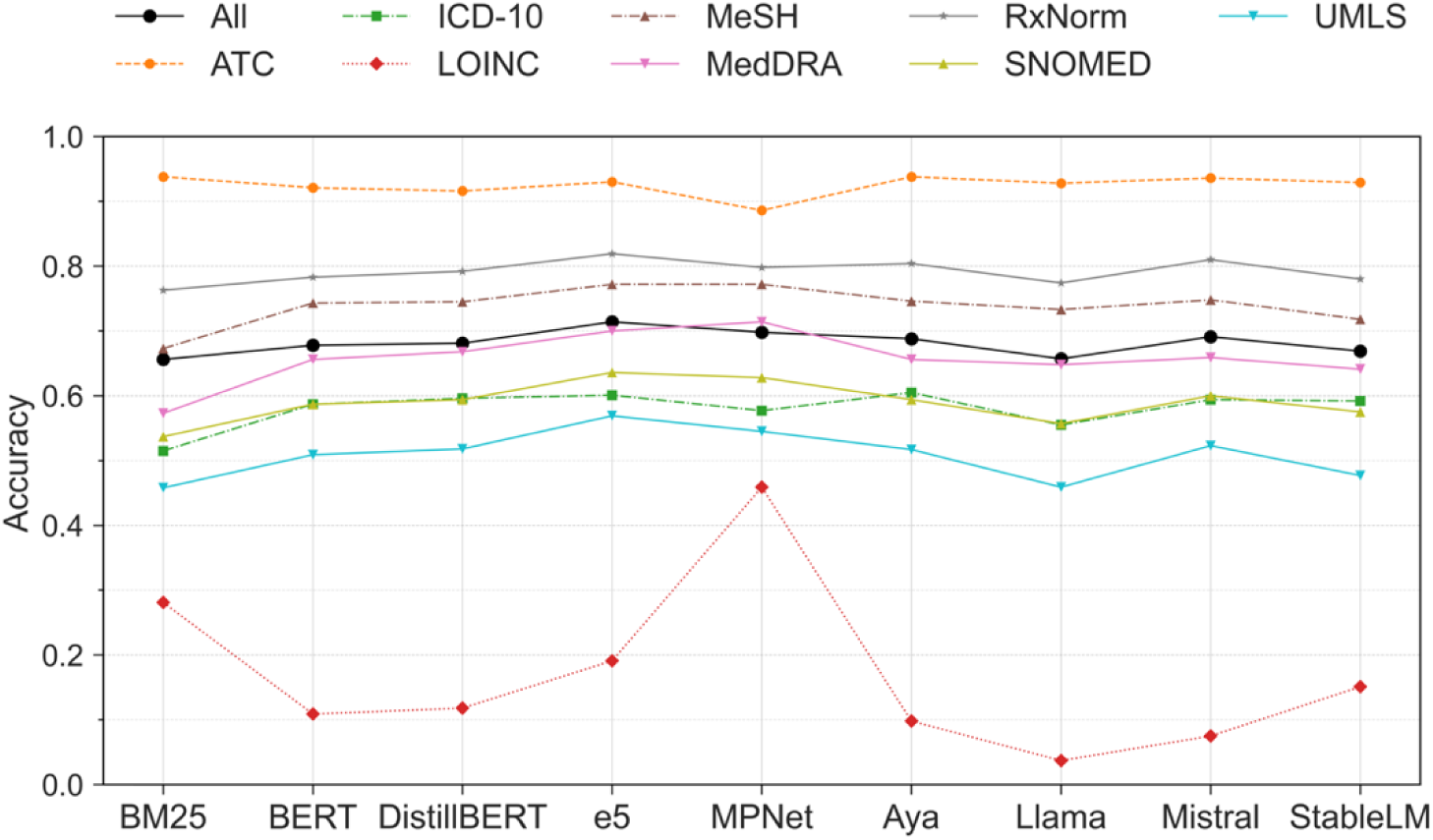
Performance comparison of LLMs on the MBCN dataset, divided by source knowledge organization system.

**Supplementary Figure 3-a:**
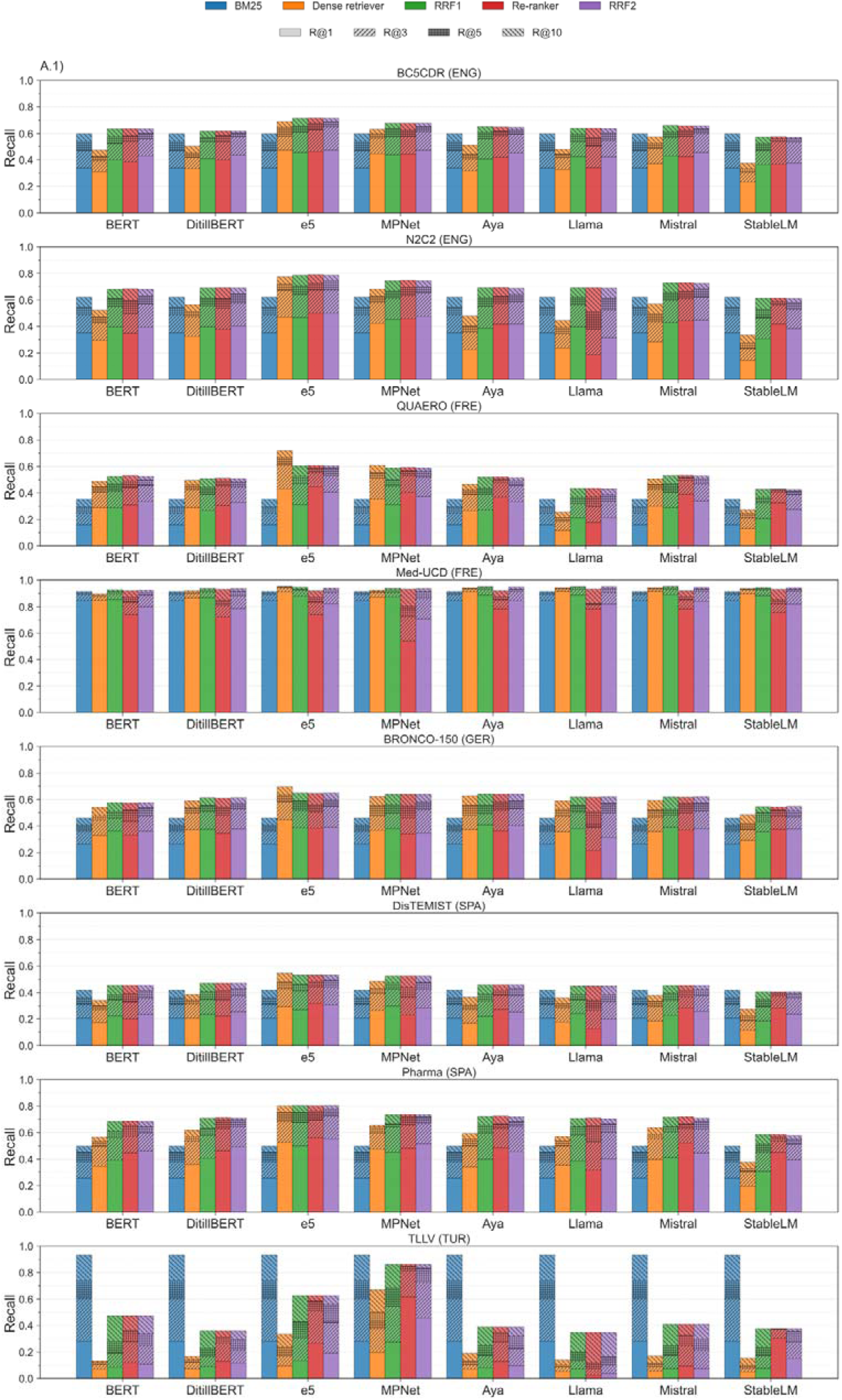
Analysis of concept normalization pipeline for different datasets, divided by steps.

**Supplementary Figure 3-b:**
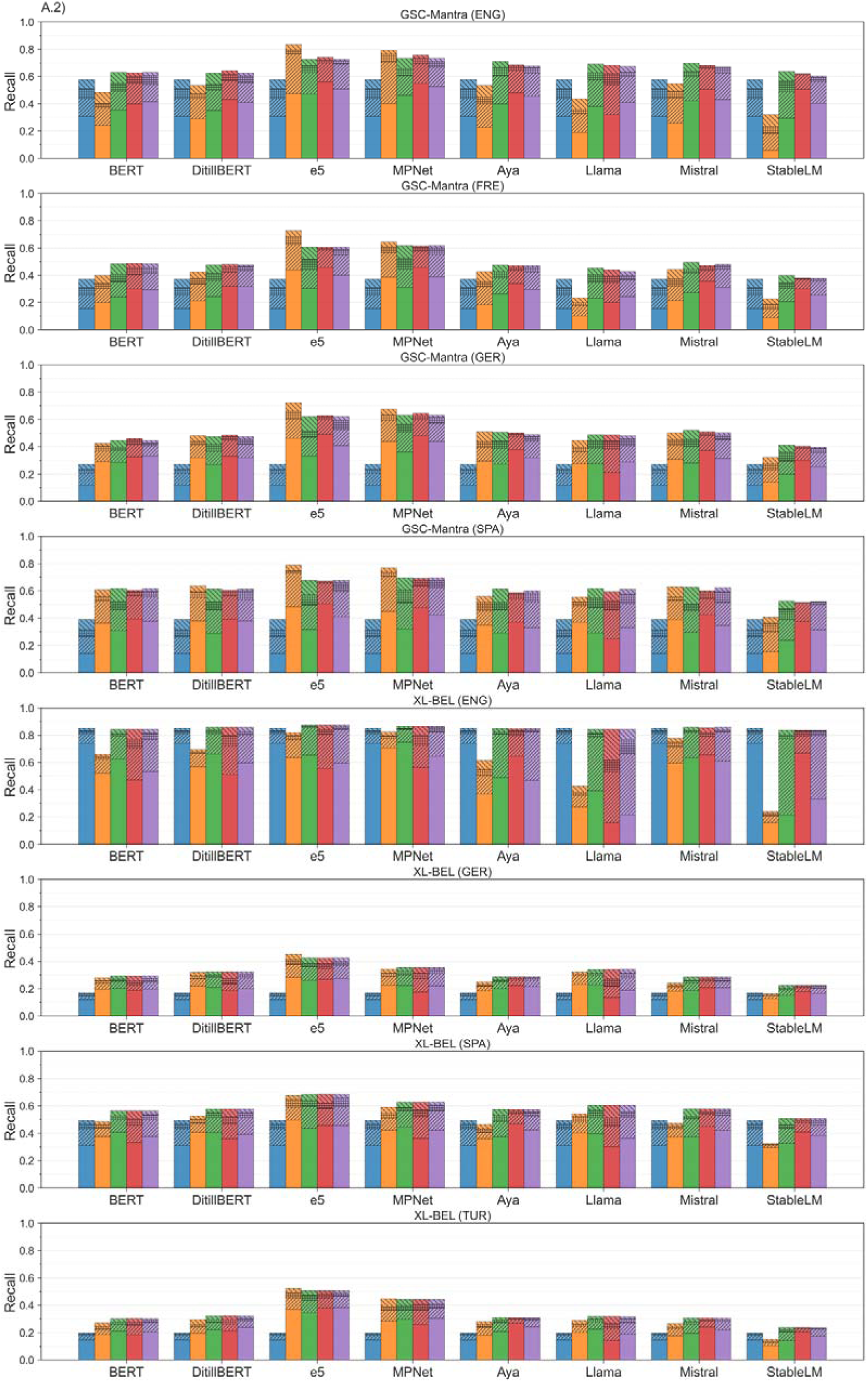
Analysis of concept normalization pipeline for different datasets, divided by steps.

**Supplementary Figure 3-c:**
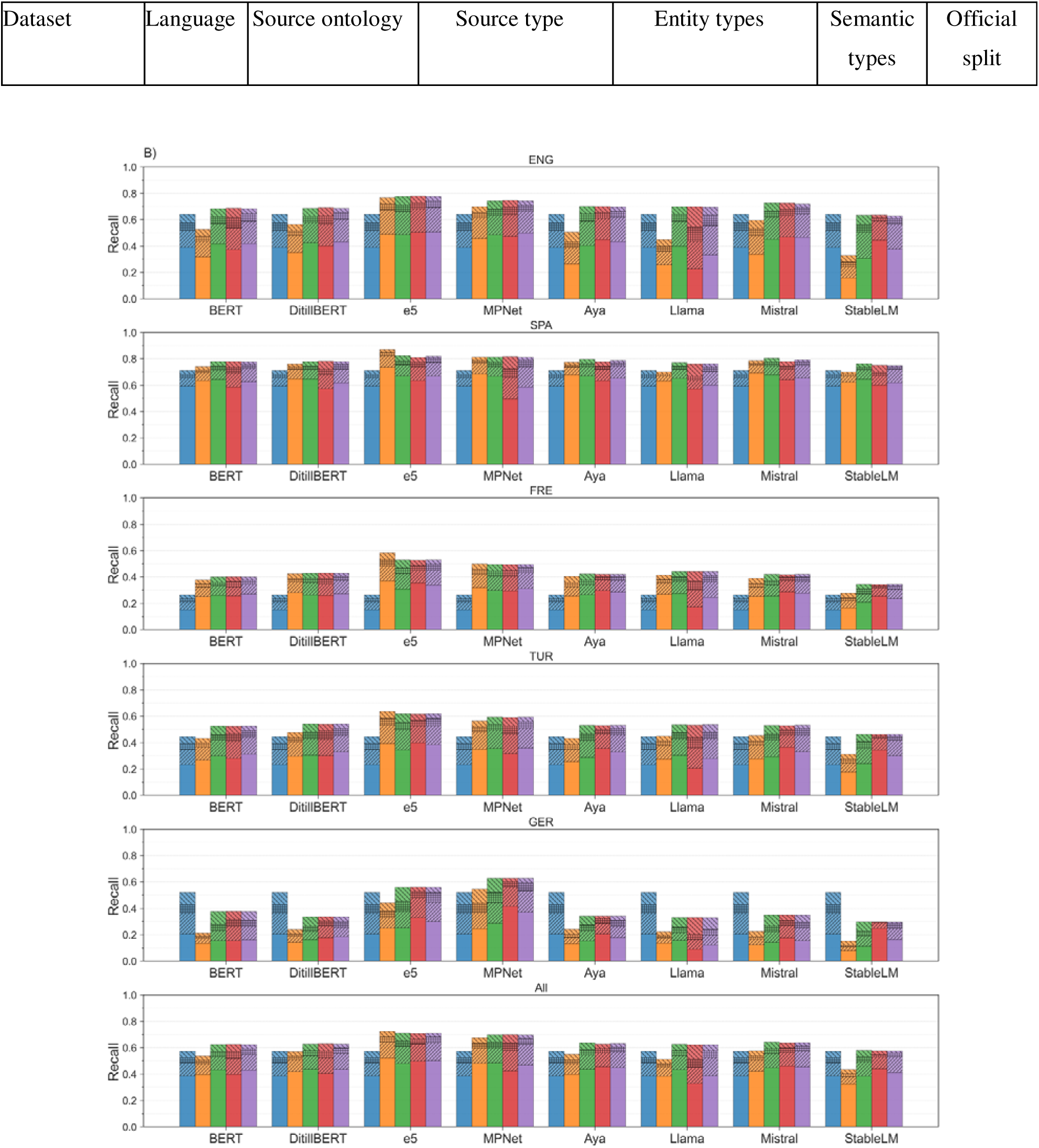
Step-by-step analysis of concept normalization pipeline for different languages.

**Supplementary Table 2.**
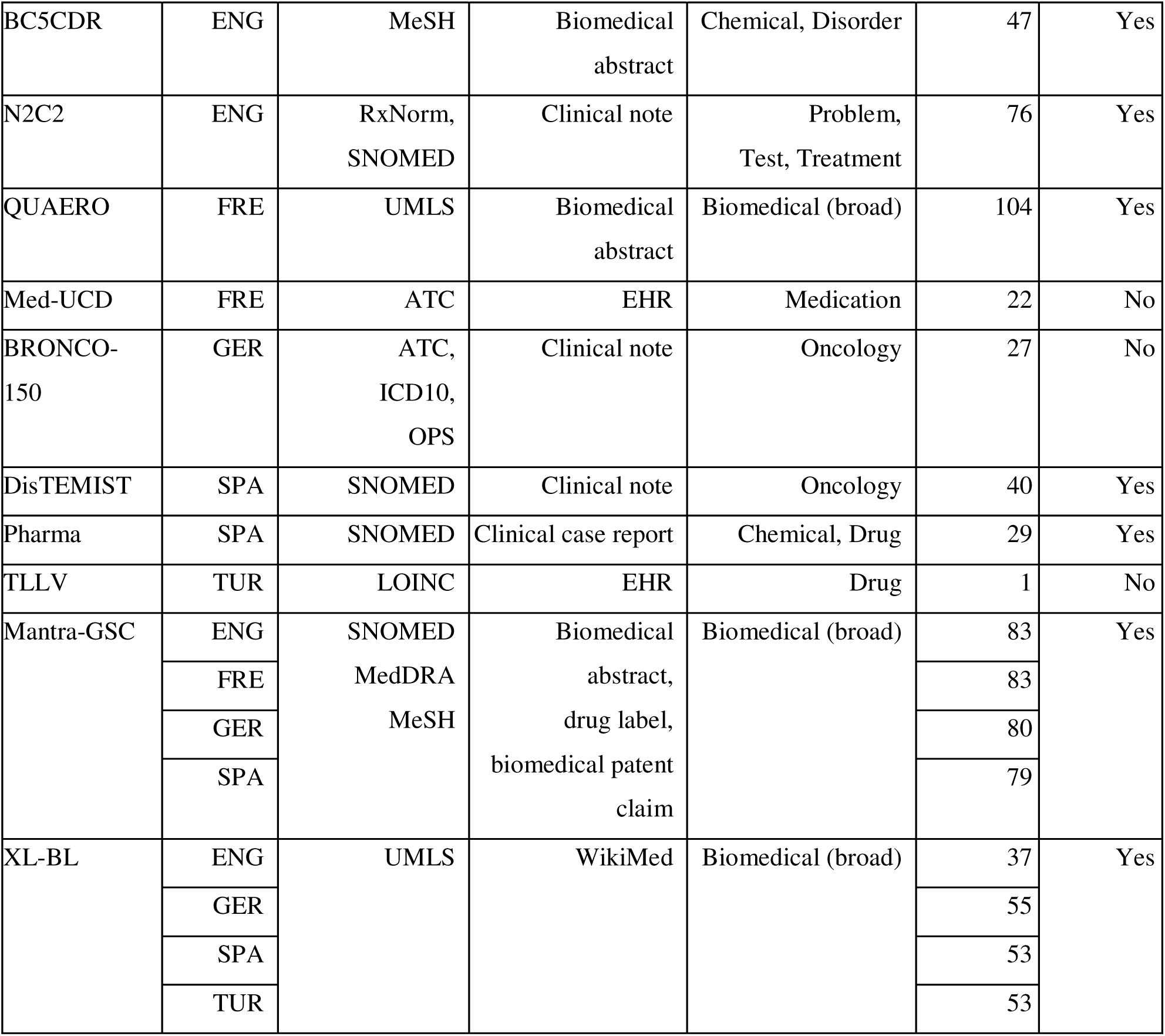
Overview of the dataset used in our experiments, including target language, target terminology/ontologies, document type, entity types, number of mentions, and unique CUIs for each dataset.

